# The Epidemiology of Scrub typhus in Thiruvarur, Tamil Nadu

**DOI:** 10.1101/2024.12.06.24317365

**Authors:** SK Farhat, M Nataraj, Sujit Kumar Behara, A Rajalakshmi, S Sweta, Sathya Jeevitha, S Binduja, S Shanti, P.K. Srivastava, Balachandar Vellingiri, Mansi Malik, Jayalakshmi Krishnan

## Abstract

National Vector-Borne Disease Control Programme (NVBDCP) under the National Center for Vector Borne Diseases Control (NCVBDC), Govt. of India, Delhi has put forth efforts to combat diseases transmitted by vectors, under the overarching umbrella of the National Health Mission (NHM). NCVBDC focuses mainly on six Vector Borne Diseases (VBD) but at the state and district level the VBD officials have to deal with all the VBDs covered under NCVBDC or outside the purview of NCVBDC. The advancement in modern technologies has increased attention to neglected tropical diseases, emphasizing their importance in public health discussions. Scrub typhus, a rickettsial infection with a harmless bite of infected chigger mite on humans, transmits the gram-negative bacteria Orientia tsutsugamushi causing the infection with rising mortality and morbidity rates across the globe. The living environment of the people plays a pivotal role in scrub typhus disease transmission. Rodents are the major reservoirs carrying the vector mites with additionally the environment, sanitation and hygiene as a crucial factor in the spread of scrub typhus.

A study was conducted from December 2023-July 2024, in the delta district of Thiruvarur covering the household participants of 730 from both rural and urban households. The statistical analysis of both quantitative and categorical variables was done using the SPSS software Version 16.0. The spacio-temporal mapping of the household areas enhanced the study with the representation of the study participants who were exposed to the risk factors but were susceptible hosts to scrub typhus. The univariate analysis showed a significant association between scrub typhus exposure with the people working in the agricultural fields, location of toilets, kitchen, presence of animals in homes, barren land and paddy fields near living areas, lack of protective implements against rodents, drying clothes on bushes, walking barefoot with more chances of being bitten by insects, mites or ticks resulting in rashes or Eschars, hospitalization due to fever, sneezing and headache. The Multivariate regression analysis showed that the association between scrub typhus exposure to the kitchen located outdoors (aOR=3.768, CI: 1.036 - 13.699, p = 0.044), people living near paddy fields (aOR=2.140, CI: 1.019-4.492, p=0.044), the use of protective implements (aOR = 0.071 (CI: 0.034 - 0.147, p < 0.001), drying clothes on bushes (aOR = 3.012 CI: 1.579 - 5.745, p = 0.001) showed strong association to exposure to scrub typhus.

## 1 Introduction

Rickettsial infections with the spotted fever group, typhus group, and scrub typhus [1] are emerging in tropical and subtropical regions but remain neglected. Despite being not included on the official list of Neglected Tropical Diseases formed by the World Health Organization’s report (WHO) [2], rickettsial diseases is ought to remain a public health concern with scrub typhus cases affecting billions of people globally with a case fatality rate of 33.3% in India [3,4]. The cases of scrub typhus in India are reported from across the nation with the Southern states of Tamil Nadu, Kerala, Andhra Pradesh, and Karnataka, Northern states of Himachal Pradesh, Jammu and Kashmir, and Uttaranchal, Eastern states of Odisha, West Bengal, and Bihar, western states of Maharashtra and northeastern states of Nagaland[5] . The infected larval chigger mites with gram-negative obligate intracellular bacteria Orientia tsutsugamushi [6] are the key vectors of scrub typhus. Being the vectors for scrub typhus, these six-legged[7] tiny chiggers are found bloated into the rodent’s and shrews’ bodies, especially to their ears, and pinnacles [8] and gets inflated as clusters on leaves, grasses, twigs and on the soil surface [7] . The Leptotrombidium deliense species of chigger mites are the main vectors causing scrub typhus in India and only chiggers of these Leptotrombidium genera are known to carry the bacteria, thus transmitting the disease to humans as accidental hosts[9], except in Chile, where they have found the efficiency of Herpetacarus antarctica carrying Candidatus Orientia chiloensisas, a vector for scrub typhus [10]. The disease with its unified sign of Eschar, with other common febrile symptoms are categorized under Pyrexia of Unknown Origin (PUO) with an undifferentiated and under-diagnosed illness[11–13]

For an illness to burn-up in the population, the risk factors act as a precursor by fueling up the disease. Thus, to study the in-depth on the infectious disease epidemiology, it is important to study the epidemiological triad [14] on the agent, host and environment to know about the disease transmission. The disease mainly affects the population living with low socioeconomic rural inhabitants with limited access to health care facilities[15]. The people living with exposure to poor hygiene and sanitation, limited access to healthcare facilities, and proximity to man-animal contact lead to the raise in prevalence of scrub typhus cases associated with the risk factors[16] . It is important to highlight the risk factors associated to scrub typhus and adopt a comprehensive approach for individuals vulnerable to all vector-borne diseases to lower the growing incidence of disease mortality and morbidity. The synanthropic exposure of humans to rodents bring individuals into close contact with chigger mites, and household surveys offer valuable insights into the environmental risks, socioeconomic status, and living conditions of participants susceptible to the targeted diseases[17,18].

Considering the above background, our study was conducted from December 2023-July 2024, in the delta district of Thiruvarur, with an objective of studying the exposure of risk factors from the case reported blocks with covering the household participants of 730 (with the mean age of 36-46 years) from both rural and urban areas. The statistical analysis of both quantitative and categorical variables were analyzed using the SPSS software Version 16.0. The spacio-temporal mapping of the household areas enabled us to emphasize on the representation about the various risk factors exposed by the study participants and were susceptible hosts to scrub typhus.

## 2 Materials and methods

This descriptive cross-sectional study was conducted from December 2023-July 2024 to access the exposure of human participants to scrub typhus in the delta district of Thiruvarur, Tamil Nadu.

### 2.1 Sampling

With mapping of secondary data of scrub typhus cases issued by the Directorate of Public Health (DPH), Chennai, out of the ten blocks of Thiruvarur district, five sampling blocks reporting highest number of cases of scrub typhus from 2018-2023 were included in our study **Fig 1**. The study sites were divided into different zones namely, North (Nannilam, Koradachery), South (Mannargudi), East (Thiruvarur) and West (Nedamangalam) shown in **Fig 2**.

**Fig 1:**
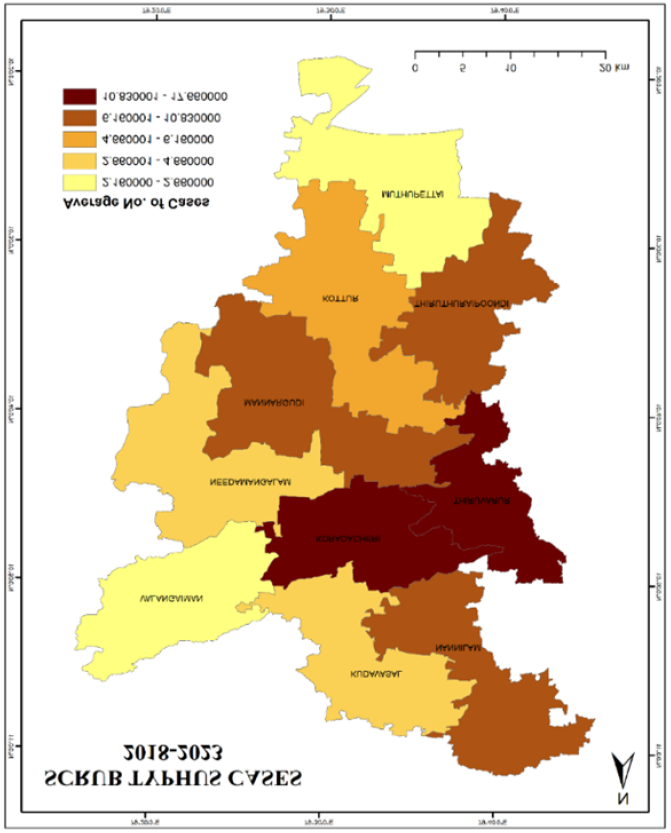
Average number of Scrub typhus cases of Thiruvarur district 2018-2023

**Fig 2:**
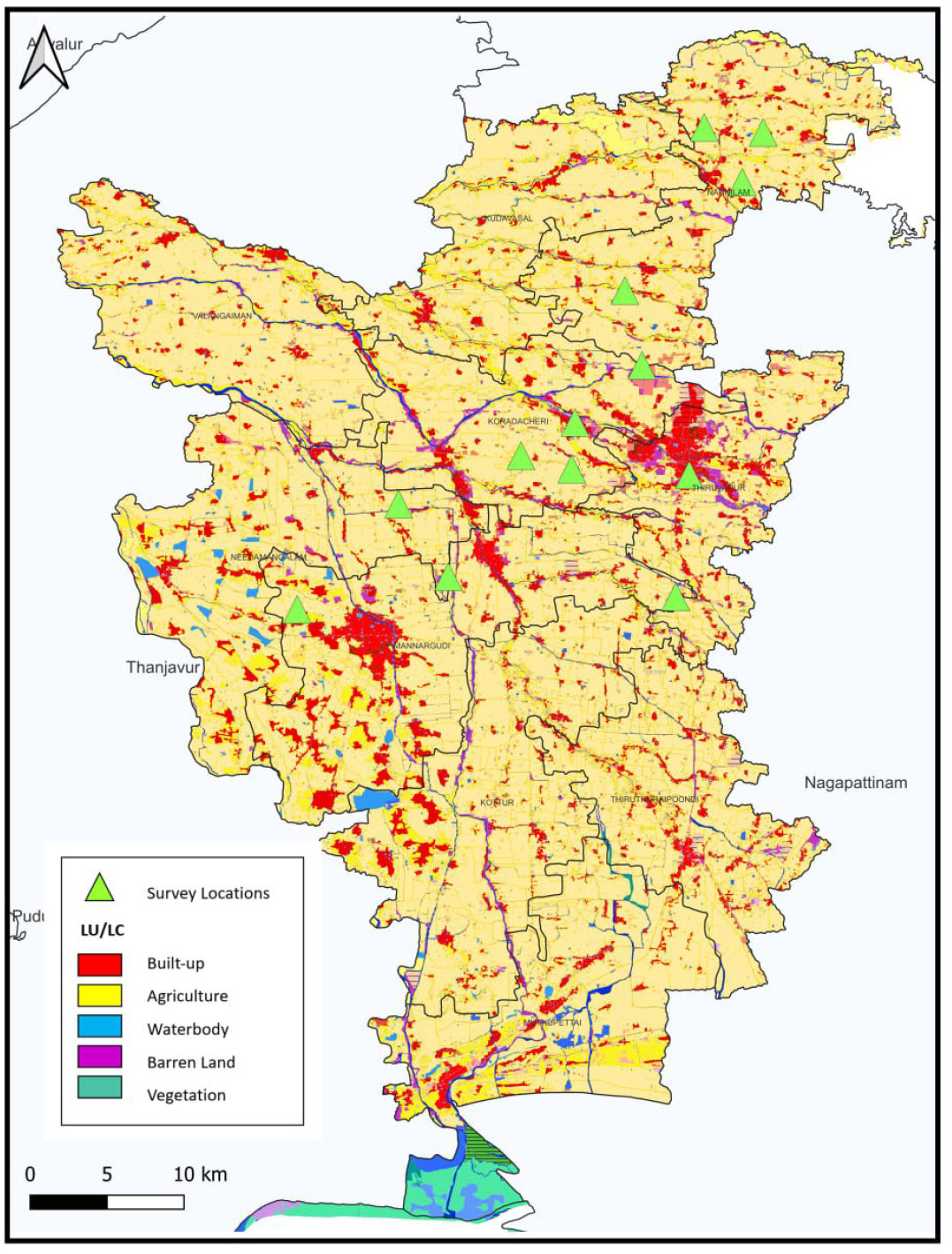
Map showcasing the study locations surrounded by type of lands

We designed a cross-sectional population survey with cluster sampling design. The sample size 730 was estimated using the STATCALC version 7.2.5.0 considering the sero prevalence of scrub typhus in India to be around 31.8% [16] with margin of error as 5%, and design effect of 2 % [19]

Fourteen clusters were chosen and within each cluster, the houses were chosen randomly using random number generator. Within a household, one participant was chosen randomly at the time of the visit from December 2023-July 2024. A total of 50-70 participants were recruited from each cluster to get the final sample size of 730.

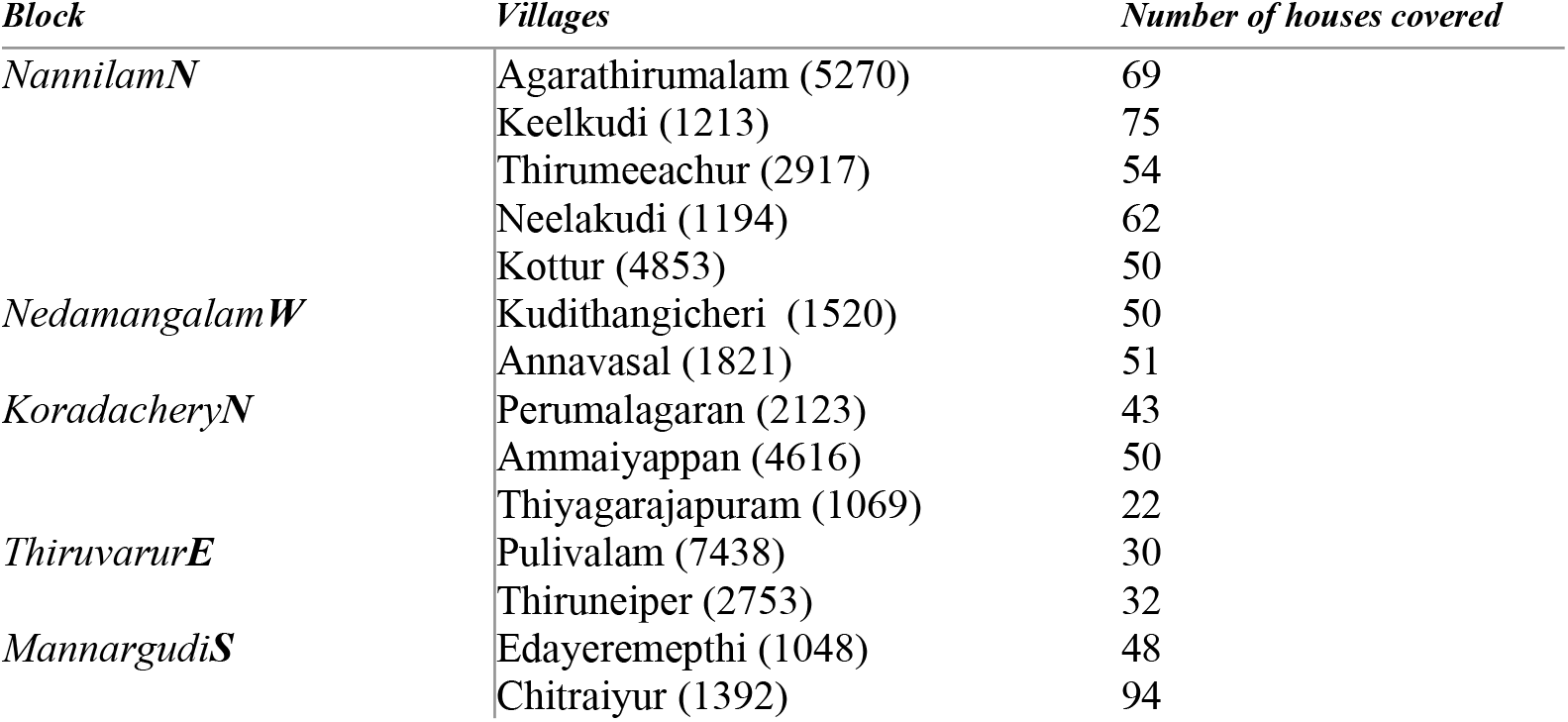

### 2.2 Data collection

A structured questionnaire format encompassing on exposure factors to scrub typhus among the chosen vulnerable human participants was developed. An information sheet with the study objectives and the consent form was distributed to the participants, choosing each participant from each household. The participants with informed consent filled out the questionnaire within 10-15 minutes. The illiterate participants were assisted by the researcher by dictating the questions and filling out the questionnaire based on their response. The participants of the age group above 12 were only included in our study.

### 2.3 Variables of the Questionnaire

The variables exposing to the risk factors for scrub typhus were included in the household survey. The questionnaire was divided into four parts with part I covering the “General information of the participants”, part II covering the “Assessment of residential area”, part III covering the “Occupational exposure”, part IV covering the “Health status of the participants” such as cardiorespiratory discomfort, and other respiratory illnesses if any. The validity of the questionnaire was tested by employing a pilot study in the sampling sites before rolling out the survey. Those respondents participated in the pilot study were excluded during final survey.

#### Dependent variable

The study used the exposure to scrub typhus as a dependent variable to study about how it is being affected by the independent variables.

#### Independent variables

The independent variables used in this study were the socioeconomic factors, housing and infrastructure, environmental sanitation and hygiene, health and demographic factors, divided into different parts.

### 2.4 Statistical Analysis

The data was entered into the Excel sheet and was imported to the IBM statistical software, SPSS (Version 16.0), an analysis including the socio-demographic variables with their association to risk factors were carried out using the chi-square analysis.

### 2.5 Ethics Statement

The study was approved by the Institutional Human Ethics Review Board **(IHERB No: CUTN/IHERB/2023-045 R1)** and the Director of Public Health & Preventive Medicine Chennai **(R.No.011575/HEB/A2/2023)** for carrying out the household survey on selected blocks of the Thiruvarur district. A verbal consent was taken from the parents/guardians for those children below 18 years for their participation in the study.

## 3.0 Results

### 3.1 Characteristics of Household Participants

A total of 730 participants were enrolled in our study by filling up the questionnaire. The majority of the participants were females 455 (62.3%) of the age group 41-60 years 276 (37.8%). The education was fairly balanced among the participants with 52.2% having primary education and 47.8% having secondary education. The study was conducted among the participants residing in the blocks of Thiruvarur district that reported scrub typhus cases from the past five years. The targeted village blocks (Nannilam, Nedamangalam, Koradachery, Thiruvarur, and Mannargudi) were further grouped into North, South, East, and West directions using the ArcGIS 10.4 software. Upon analyzing the general characters of the participants, **Table 1**, it was noted that over half of the population lives in Kutcha houses (55.3%), and most have outdoor toilets (82.1%) and indoor kitchens (87.5%). A significant portion of the population (58.4%) lived with animals, and 84.2% resided near barren land, primarily paddy fields (60.3%). Scrub typhus exposure showed affecting 76.8% of the population, yet only 39.3% had protective measures in place. Half of the population walked barefoot, and 60% worked in agriculture or forestry-related fields. Health issues were common, with 28.1% reporting fever in the past month and 31.1% experiencing breathlessness, while conditions like body aches, chest discomfort, and eye irritation were also prevalent. Despite frequent illnesses, only 3.8% were hospitalized due to fever. Overall, the data reflects a population with significant environmental exposure and health challenges, particularly exposure to scrub typhus and other outdoor living conditions.

**Table 1:**
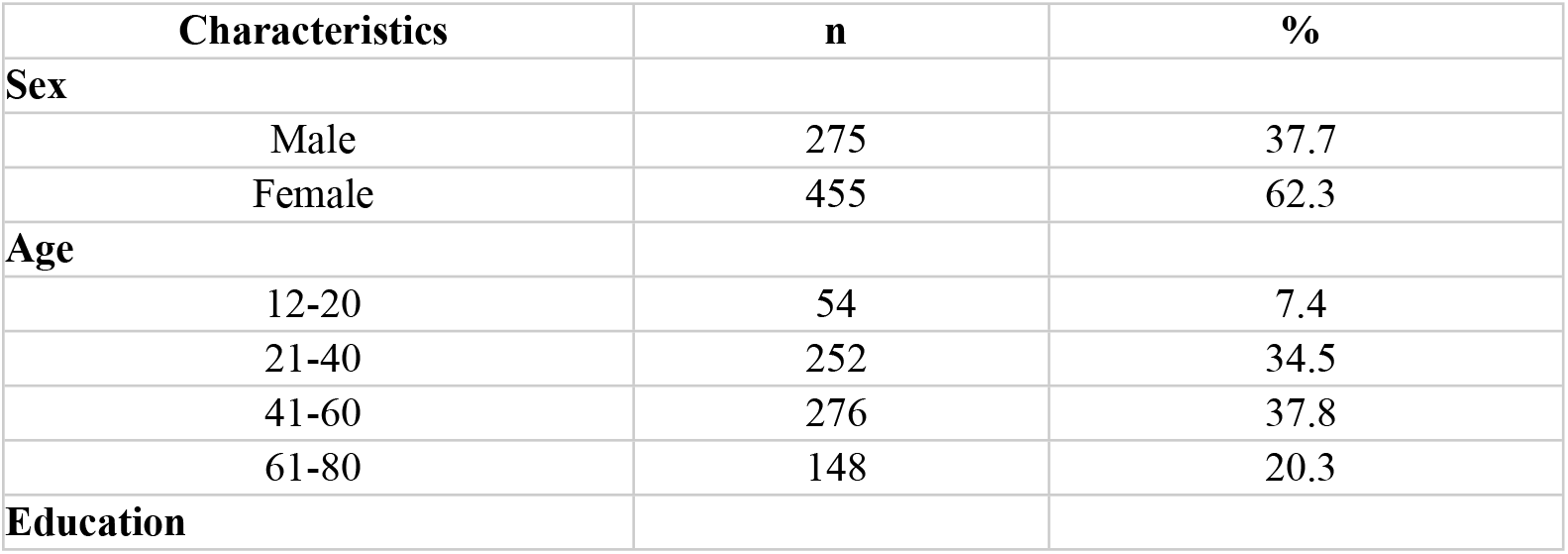
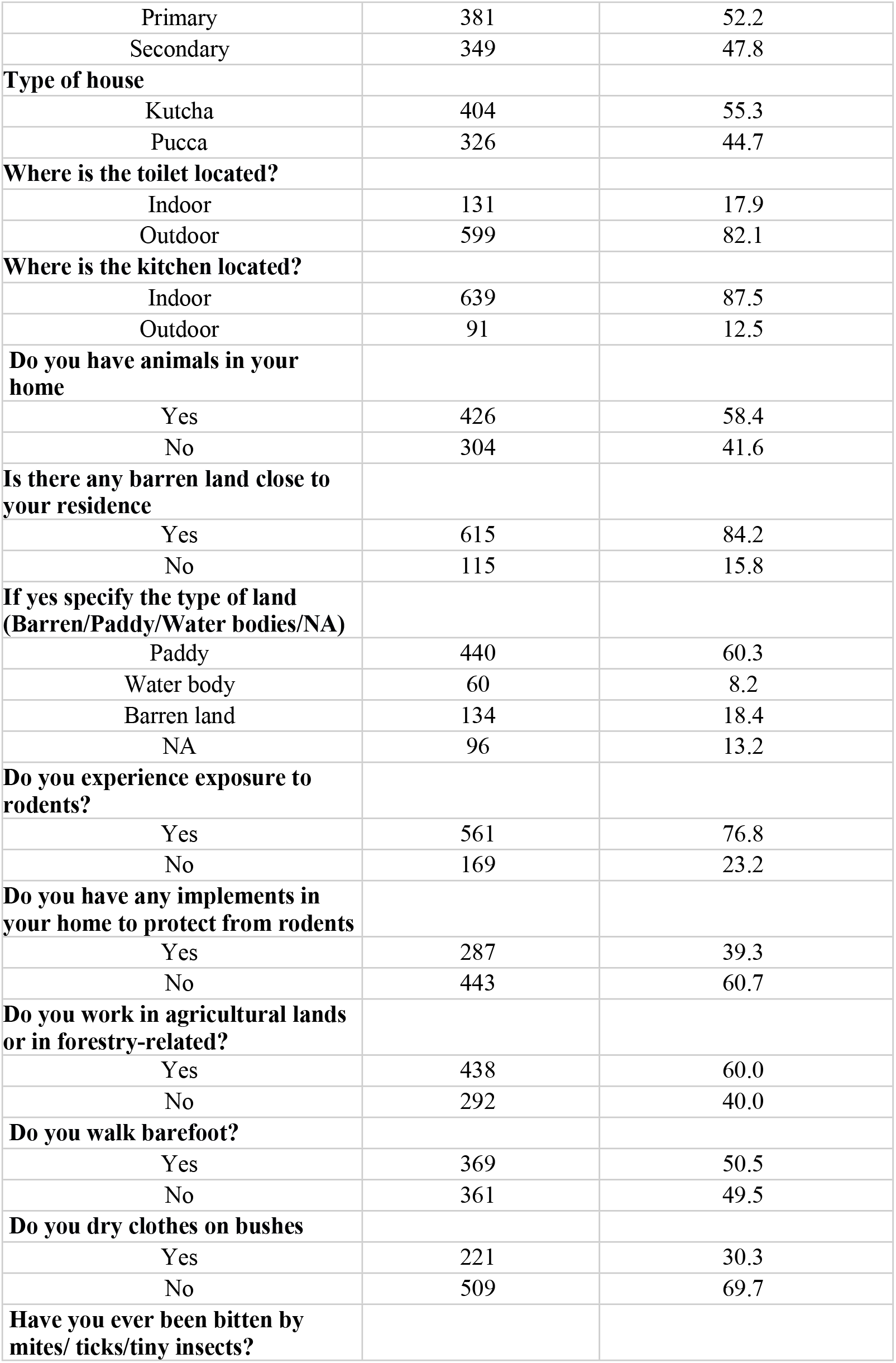
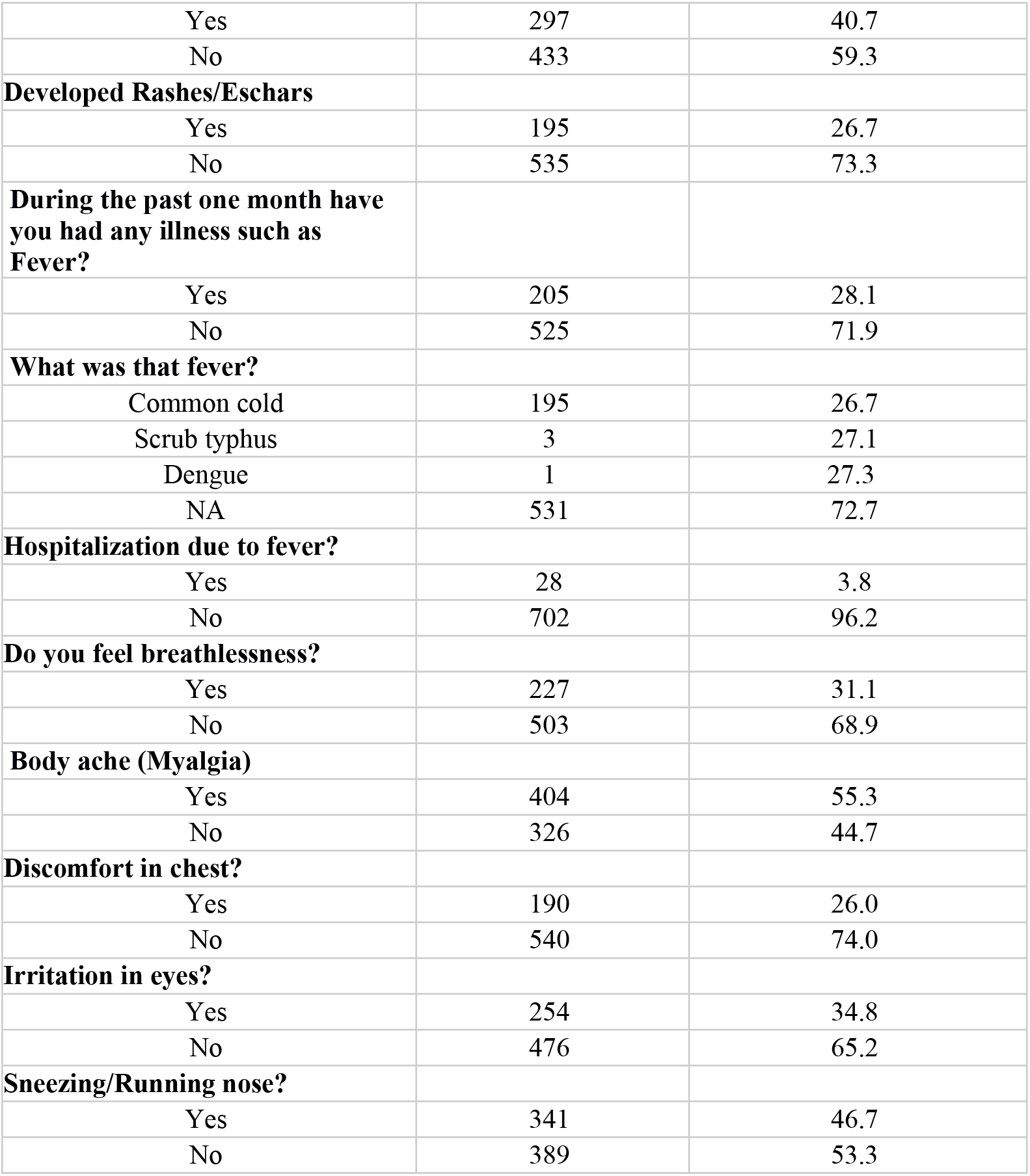
General Characteristics of Household Participants.

### 3.2 Univariate Analysis of study participants

**Table 2**, depicts the Univariate analysis with a significant difference noted for the variables including location of toilet with 78.8% people using outdoor toilets with exposure to scrub typhus, compared to indoor toilets (p=0.008). The outdoor location of the kitchen in the households (96.7%) showed statistically significant difference to the exposure of scrub typhus (p=<0.001). The presence of animals in the home (79.6%) showed significant exposure to scrub typhus with (p=0.039). The presence of barren land across the living habitats of the participants (78.5%) showed more exposure to scrub typhus with (p=0.012) showing statistical significance with paddy fields showing the highest exposure of scrub typhus (85%), followed by the presence of water bodies (73.3%) and barren lands (61.2%) close to the proximity (p<0.001). Upon investigating on the usage of any rodent protective implements, it was noted that those using rodent protections such as traps showed significant association to exposure to scrub typhus (p<0.001). The practice of people drying clothes on bushes (93.2%) showed more exposure to scrub typhus (p<0.001) compared to those who do not dry clothes on bushes (69.7%). Walking barefoot participants, showed significant exposure to scrub typhus (84.6%) (p<0.001) compared to those who use protective measures. The people working in the fields, as agriculturalists, showed significant strong association to exposure to scrub typhus (81.1%) with those non-workers (70.5%) (p=0.001). The participants exposed to rodents had reported history of insect, mite or tick bite (84.8%) (p<0.001). The health-related symptoms such as sneezing/running nose (p<0.001), headache (p=0.002), and development of eschars/rashes (p=0.001) showed significant association to scrub typhus. A borderline significant difference was noted in this study with high exposure of rodent in participants hospitalized due to fever (85.7%) (p=0.049). However, no statistical significance was seen between type of house, household size, education, sex and other health conditions such as fever, body aches, chest discomfort and breathlessness.

**Table 2:**
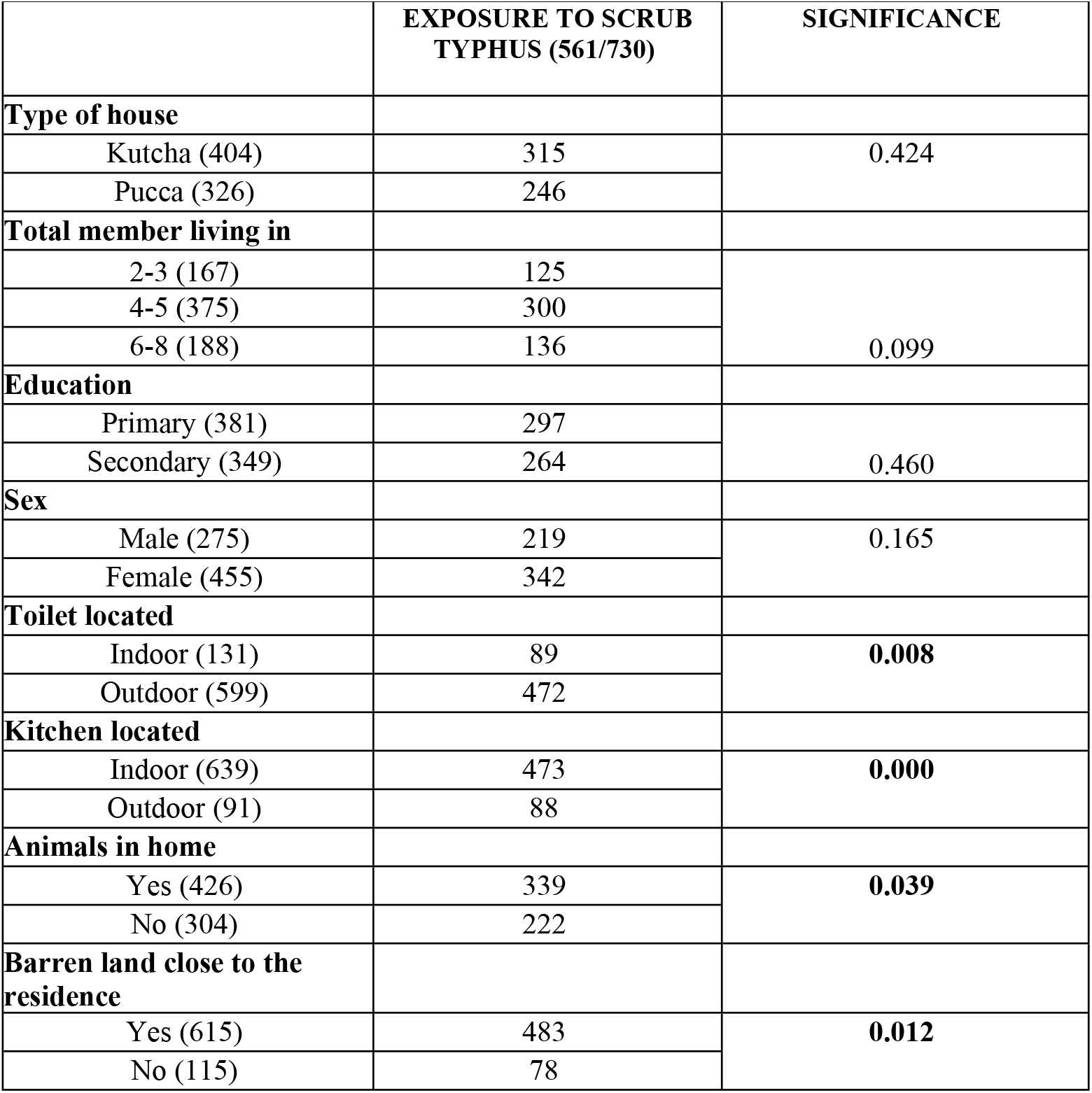
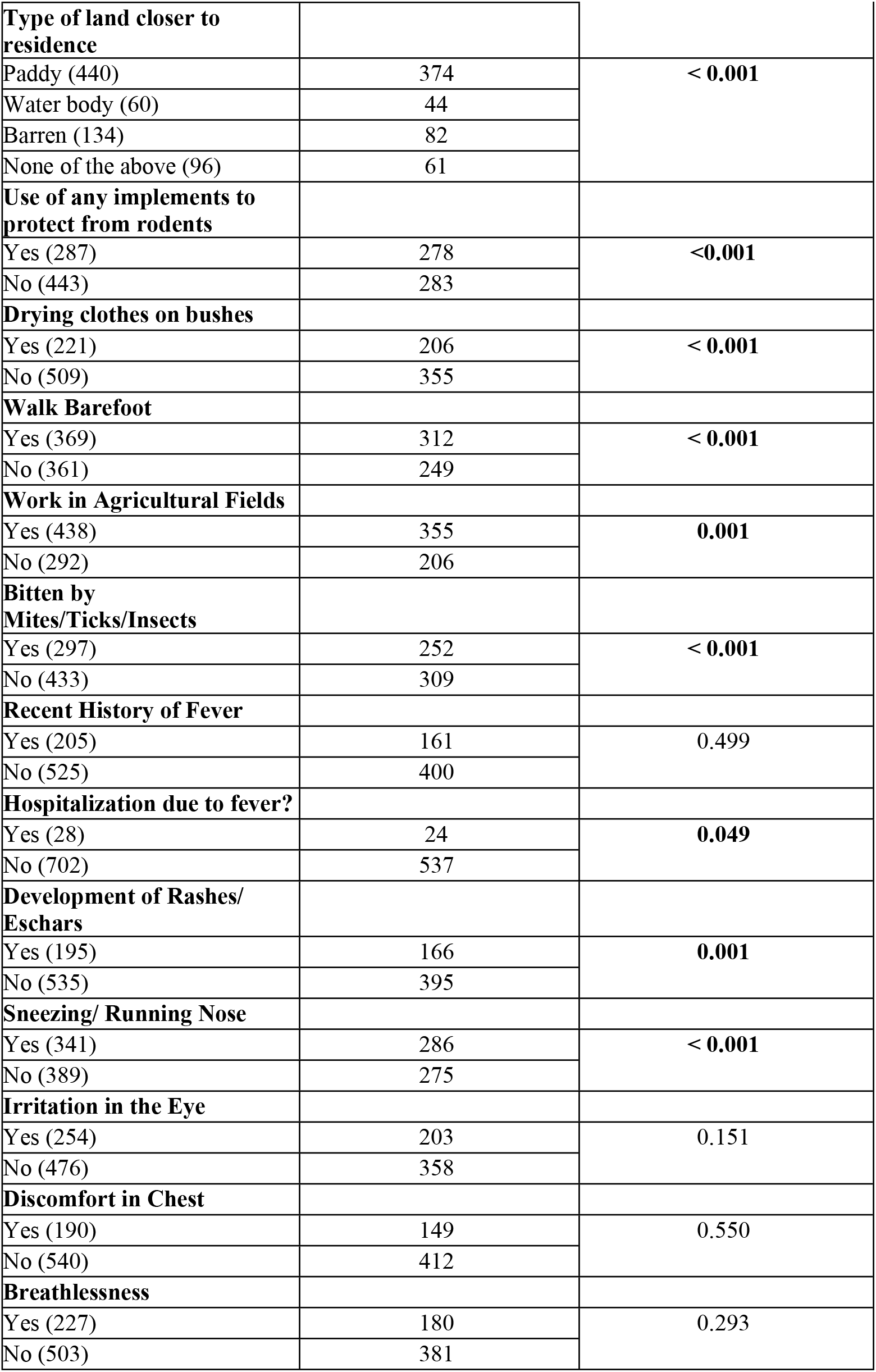
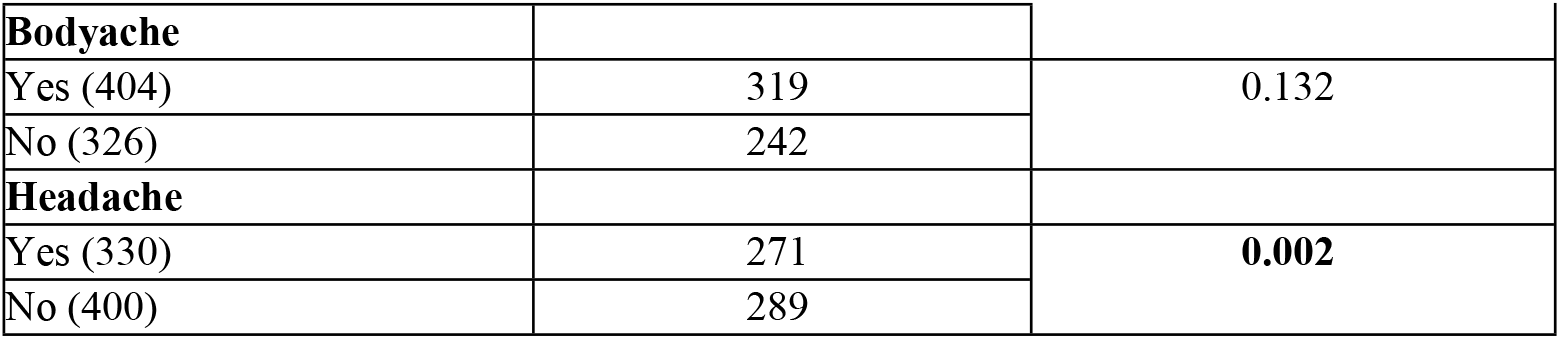
Univariate Analysis of Household Participants.

### 3.3 Multivariate Analysis

The statistically significant variables in the Univariate analysis such as the location of toilet and kitchen at home, presence of animals as pets at home, the type of barren land close to the residence, the usage of any implements to protect from rodents, the practice of drying clothes on bushes and walking barefoot with working in the fields of agriculture, often getting bitten by mites, ticks and insects, hospitalization due to fever, development of rashes or eschars, sneezing or runny nose and headache were included for further analysis in multiple regression analysis since they showed low co-linearity.

The results showed a markedly significant association between the dependent variable of exposure to scrub typhus, and the independent variables. The odds of having an exposure to scrub typhus was found to be 3.768 times greater (aOR=3.768, CI: 1.036 - 13.699, p = 0.044) in those households where kitchen was located outdoors compared to kitchen located inside. The odds of having exposure to scrub typhus (aOR=2.140, CI: 1.019-4.492, p=0.044) among people living near paddy fields showed statistical significance **Fig 3**. The people living near water bodies did not significantly affect exposure to scrub typhus (p > 0.05). The protective implements with (aOR = 0.071 (CI: 0.034 - 0.147, p = 0.000) were strongly associated with lower exposure of rodents (highly significant, p < 0.001). Drying clothes on bushes (aOR = 3.012 CI: 1.579 - 5.745, p = 0.001) showed significant increase for the exposure to scrub typhus (p = 0.001).

| Variables |  | Unadjusted odds ratios (uOR) |  | Adjusted odds ratios (aOR) |  |  |  |
| --- | --- | --- | --- | --- | --- | --- | --- |
|  |  | Exp(β) | p-value* | Exp(β) | 95%ConfidenceInterval |  | p-value* |
|  |  |  |  |  | LowerBound | UpperBound |  |
| **Toilet located** | Outdoor | .985 | .938 | 1.375 | 0.832 | 2.271 | 0.214 |
|  | Indoor (Ref) | 1 |  | 1 |  |  |  |
| **Kitchen located** | Outdoor | .285 | **.000** | 3.768 | 1.036 | 13.699 | **.044\*** |
|  | Indoor (Ref) | 1 |  | 1 |  |  |  |
| **Animals in home** | Yes | .746 | .053 | .887 | .592 | 1.329 | .561 |
|  | No (Ref) | 1 |  | 1 |  |  |  |
| **Barren land close to the residence** | Yes | 1.036 | .861 | .947 | .259 | 3.456 | .934 |
|  | No (Ref) | 1 |  | 1 |  |  |  |
| **Type of land closer to residence** |  |  |  |  |  |  |  |
|  | Paddy | 1.554 | 0.191 | 2.140 | 1.019 | 4.492 | **.044** |
|  | Water body | 0.726 | 0.238 | .647 | .168 | 2.496 | .527 |
|  | Barren | 1 |  | 1 |  |  |  |
| **Use of any implements to protect from rodents** | Yes | 1.824 | **.000** | .071 | .034 | .147 | **.000** |
|  | No (Ref) | 1 |  | 1 |  |  |  |
| **Drying clothes on bushes** | Yes | .551 | **.000** | 3.012 | 1.579 | 5.745 | **.001** |
|  | No (Ref) | 1 |  | 1 |  |  |  |
| **Walk Barefoot** | Yes | 1.732 | **.000** | .702 | .462 | 1.069 | .099 |
|  | No (Ref) | 1 |  | 1 |  |  |  |
| **Work in Agricultural Fields** | Yes | 1.396 | **.029** | .898 | .581 | 1.388 | .627 |
|  | No (Ref) | 1 |  | 1 |  |  |  |
| **Bitten by Mites/Ticks/Insect** | Yes | 1.326 | .062 | .723 | .451 | 1.160 | .179 |
|  | No (Ref) | 1 |  | 1 |  |  |  |
| **Hospitalization due to fever?** | Yes | .840 | .146 | 1.183 | 1.183 | .656 | .576 |
|  | No (Ref) | 1 |  | 1 |  |  |  |
| **Development of Rashes/ Eschars** | Yes | .609 | **.003** | .904 | .530 | 1.543 | .711 |
|  | No (Ref) | 1 |  | 1 |  |  |  |
| **Sneezing/ Running Nose** | Yes | .830 | .212 | 1.389 | .906 | 2.129 | .132 |
|  | No (Ref) | 1 |  | 1 |  |  |  |
| **Headache** | Yes | 1.205 | **0.012** | 1.277 | .838 | 1.945 | .255 |
|  | No (Ref) | 1 |  | 1 |  |  |  |

**Fig 3:**
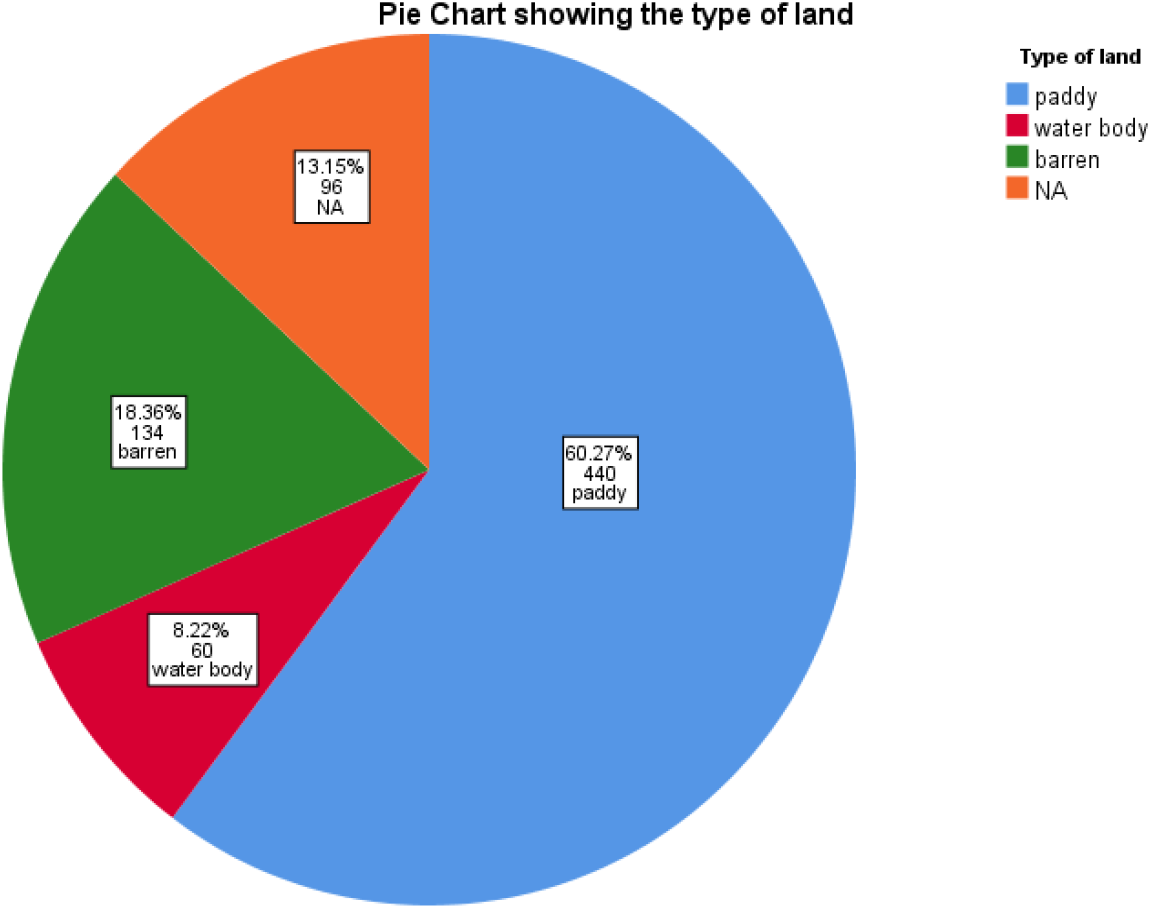
Pie chart showing the type of land occupied near to the living habitats of participants

## 4.0 Discussion

The study emphasizing on population concerning the risk factors associated with scrub typhus, as a measure of preventive health approach was conducted among the 730 household participants of the Thiruvarur district using a questionnaire. Thiruvarur is known to be the “rice bowl of Tamil Nadu” with paddy fields covering many plains, of which 70% of the population is occupied in agriculture[20] were taken into consideration. Scrub typhus is a growing concern in the district of Thiruvarur with the number of reported cases to be 121 (DDHS, Thiruvarur) as of 2021, with the hotspots of areas in the district. The occurrence of disease in the district is quite prevalent with cases increasing every year. Henceforth, with an increasing trend of clinical cases of scrub typhus in the district, it was evident that the district is endemic to scrub typhus with the population being vulnerable to the risk factors.

In the present study, it is reported that the majority of the participants were females 455 (62.3%) of the age group 41-60 years, with over half of the population living in the Kutcha houses (55.3%), and most having outdoor toilets (82.1%) and indoor kitchens (87.5%). A significant portion of the population (58.4%) lived with animals, and 84.2% and 60.3% resided near barren land, paddy fields respectively. Rodent exposure affected 76.8% of the population, yet only 39.3% had protective measures. Half of the population walked barefoot, and 60% worked in agriculture or forestry-related fields. Health issues were common, with 28.1% reporting fever in the past month and 31.1% experiencing breathlessness, while conditions like body aches, chest discomfort, and eye irritation were also reported. Despite frequent illnesses, only 3.8% were hospitalized due to fever.

The Multivariate regression analysis showed that the association between rodent exposure to the kitchen located outdoors (aOR=3.768, CI: 1.036 - 13.699, p = 0.044), people living near paddy fields (aOR=2.140, CI: 1.019-4.492, p=0.044), the use of protective implements (aOR = 0.071 (CI: 0.034 - 0.147, p < 0.001), drying clothes on bushes (aOR = 3.012 CI: 1.579 - 5.745, p = 0.001) showed strong association to exposure to rodents.

The male population of Taiwan belonging to the age group of 40-69 years old with an occupation in the outdoors and military personnel were associated with a high incidence of scrub typhus disease, with more than 50% of cases reported [21].

A study conducted in Uttar Pradesh; India showed that children exposed to an outdoor environment with open defecation, cattle contact, indoor storing of firewood, and agricultural activities showed a higher risk of acquiring the disease[22]. Open defecation in a squatting position is the main leading etiology to scrub typhus infection-causing Eschar in the perennial area of the targeted accidental human hosts [23]. Considering the emerging cases of scrub typhus as a public health threat, a study at Vellore, on accessing the risk factors showed that the majority of the cases were the agricultural laborers living close to bushes and shrubs, with single-roomed homes [24]. Unstructured occupational health practices among people working in agricultural fields, especially in paddy and vegetable farms with exposure to scrub typhus, and domestic animals showed significant risk factors of having scrub typhus[25] . Occupational exposure in the agricultural fields, animal husbandry, and armed personnel showed a significant association with scrub typhus cases in Taiwan [21]. The household participants with a nuisance to mice in their home, and with poor sanitation showed significant association to acquiring scrub typhus infection[26]. People working in croplands, with outdoor toiletries and pets, and sitting on grasses showed significant risk factors to developing scrub typhus (odds ratio 1,519; p<0.001) [27].

Our study showed results consistent with those of [28], who found a significant association between the location of the kitchen in outdoors to an increased risk of acquiring scrub typhus (OR 5.61; 95% CI: 1.51–23.01). Similarly, our findings also revealed a significant association between location of kitchens outdoor to exposure to scrub typhus (aOR = 3.768; 95% CI: 1.036–13.699; p = 0.044). Additionally, our study identified a significant risk associated with drying clothes on bushes (aOR = 3.012; 95% CI: 1.579–5.745; p = 0.001), further increasing the likelihood of contracting the disease. In contrast, drying clothes on a clothesline, as reported by [28], demonstrated a protective effect, though it did not reach statistical significance (OR 0.31; 95% CI: 0.08–1.08; p = 0.077). Furthermore, the proximity of paddy fields to households was associated with a greater risk of scrub typhus (aOR = 2.140; 95% CI: 1.019–4.492; p = 0.044), compared to areas near water bodies. The use of protective measures significantly reduced exposure to scrub typhus (aOR = 0.071; 95% CI: 0.034–0.147; p < 0.001), as highlighted by [5], suggesting these measures are an effective means of controlling scrub typhus.

Although the basic epidemiological characters like the demography, occupation, and environment were focused on the research studies conducted to study the etiology of scrub typhus [21,29], other factors like the temperature and rainfall were also found to be the contributing risk factors for scrub typhus [30] . Deforestation is also a leading cause of risk factor for scrub typhus with people being more exposed to rodents and animals with increasing cases of incidence of scrub typhus in South Korea [31] .

The emerging trend of neglected tropical diseases has now put forth the limelight on their control and prevention. The various national health programs for communicable and non-communicable diseases by the government have strengthened the seriousness and ill effects of vector-borne diseases in the population. The active screening and surveillance of vectors with an aim of source reduction is the target to control and prevent vector-borne diseases. Eliminating the risk factors contributing to the disease at its primordial prevention will lower the disease burden. The policy makers along with the public health professionals should comprehensively evaluate the disease incidence and timely amendment of the programs as its necessity is required.

The basic knowledge about the disease scrub typhus and its transmission was assimilated by the study participants with the concept of health promotion and primordial disease prevention. With focusing on the impact of disease severity, the study participants were taught through ICT (Information and Communication Technologies) about basic sanitation and hygiene practices and to control rodent exposure in households.

## 5.0 Conclusion

The study reveals a statistical association between the risk factors and their impact on rising cases of scrub typhus across the globe thus flagging that it is a public health concern to control the disease at community level through individual approach. The need of having a National Programme focussing to control the rodent reservoirs carrying the vectors of scrub typhus and its vector control activities is of utmost importance. The appropriate vector control strategy such as using pyrethrum or allethrin aerosol bombs, rotenone or pyrethrum sprays, wiping the woods or furniture with cloth saturated with kerosene or any alternatives as kerosine is not available everywhere, and vacuum cleaners etc., is also crucial.

Information on scrub typhus, Education and Communication (IEC) and Communication Behavioral Impact (COMBI) are the most appropriate tools, as the people of the village still do practice open defecation, do dry their clothes on the bushes, walk bare foot as there is lack of awareness about the disease severity and its transmission.

An Eschar, which is the main symptom of scrub typhus must be diagnosed by the physicians early as they are found mostly in genital areas. The history of the patients need to be recorded properly. The vector control management should be initiated to control the vectors. The primordial prevention of the disease is very important to lower the incidence of cases and it acts as a protective shield towards acquiring any illness. Therefore, studies involving survey with participants will help grab attention to the disease burden and their transmission at the primordial level than letting the disease to enter into tertiary care.

## Data Availability

All data produced in the present study are available upon reasonable request to the authors

## CRediT authorship contribution statement

**Conceptualization**-Farhat, Jayalakshmi, Sujit

**Formal analysis**: Farhat, Binduja, Swetha, Sathya Jeevitha

**Writing– original draft:** Farhat, Sujit, Jayalakshmi

**Writing– review & editing:** Rajalakshmi, Mansi, Blachander, Shanti, Natraj, PK Srivastava, Sujit

## Acknowledgment

The authors acknowledge the invaluable contributions made by the respondents. We also extend our gratitude to the district health officials and other stakeholders for their significant support in conducting the research.

## Abbreviation

aOR: Adjusted Odds Ratio
CI: Confidence Interval
WHO: World Health Organization
PUO: Pyrexia of Unknown Origin
IHERB: Institutional Human Ethics Review Board
COMBI: Communication Behavioral Impact
IEC: Information, Education, and Communication
ICT: Information and Communication Technologies
NVBDCP: National Vector Borne Disease Control Programs

